# Identifying resilience factors for adolescent mental health with cyberbullying victimisation as a risk factor

**DOI:** 10.1101/2024.11.18.24317457

**Authors:** Aaron Kandola, Rosie Mansfield, Yvonne Kelly, Yasmin Rahman, Karmel Choi, Chris Hollis, Ellen Townsend, Praveetha Patalay

## Abstract

Promoting resilience can reduce the mental health risks of cyberbullying victimisation in adolescents.

We conducted a longitudinal cohort study with 9,969 adolescents (Millennium Cohort Study (MCS)) at ages 14-15 (baseline) and 17 (follow-up). We replicated our analyses in 4,240 adolescents (Longitudinal Study of Australian Children). The outcome was psychological distress at follow-up. Cyberbullying victimisation was a single-item question. Aim one identified modifiable resilience factors (exposures) associated with psychological distress. Aim two examined whether these resilience factors (moderators) interacted with the cyberbullying-distress association.

478/1,466 baseline variables were associated with distress after confounder adjustment (aim one). 31/478 potential resilience factors moderated the cyberbullying-distress association. 15 models replicated in the individual (n=8, e.g., happiness with friends), family and friends (n=3, e.g., sexual activities), structural (n=2, e.g., income sources), and learning environment (n=2, e.g., misbehaviour) domains.

We identified several factors for further research on developing interventions to reduce adolescent cyberbullying mental health risks.

## 1. Introduction

Mental health conditions are among the leading causes of global disability and are rising in prevalence [1], including in adolescents [2]. The onset of major mental health conditions commonly first occurs during adolescence [3–5], and early onset increases the risk of lasting psychological, physical, and social problems during adulthood [6, 7]. Preventive and early intervention approaches during adolescence could improve wellbeing and reduce the risk of these longer-term problems, but we still lack effective public health measures that focus on mitigating harms from well-established risk factors [8, 9].

Cyberbullying is becoming an increasingly salient issue due to the rise in access to online platforms [10– 12] and refers to repeated, aggressive behaviour towards an individual online, such as on social media platforms [13]. Bullying victimisation during adolescence is common [14], with one UK-based study estimating that around 36% of girls and 24% of boys aged 15 experienced bullying during the last couple of months [15]. It can increase the risk of several mental health conditions during adolescence and adulthood [15–18]. For example, some studies estimate that 25-40% of the population-level variance in depression and self-harm may be due to all forms of bullying [19, 20]. Cyberbullying and face-to-face bullying are correlated but cyberbullying may have independent and additive mental health risks in young people [21, 22]. Concerns over cyberbullying as a mental health risk factor are growing with the time young people spend on social media platforms, which are becoming more numerous and pervasive [11]. Such concerns are epitomised by the UK Government recently announcing measures to ban mobile phone use in schools citing cyberbullying as a growing concern [23], while the House of Commons commissions research on the relationship between digital device use and well-being in young people [24]. Rapid advances in artificial intelligence, virtual and extended reality, and other technologies mean that cyberbullying can occur in increasingly immersive and realistic virtual settings, perpetuating its potential future mental health risks. This study is embedded within the Digital Youth programme (https://digitalyouth.ac.uk/), whereby our results will feed into studies examining certain risk factors in greater detail and the co-development of interventions to protect young people from online harms.

Its high prevalence, ease of perpetrators’ access to victims, evolving digital technologies, and clear implications for mental health make cyberbullying a clear public health problem. Schools are inconsistent in implementing anti-bullying programmes [25], which may only have a small impact on reducing face-to-face bullying [26].It is potentially even harder to reduce cyberbullying through school-based approaches alone given the ease of access to these platforms and the lack of regulatory incentives for them to promote public health [27]. Efforts to reduce and prevent bullying must continue, but increasing adolescent resilience to the mental health impact of cyberbullying is an important complementary approach to reduce its potential harm. We refer to resilience here as the process of individuals navigating biopsychosocial resources to manage and successfully adapt to stressors or trauma that may otherwise cause psychological distress [28]. Interventions that broadly promote mental health resilience can reduce depression and anxiety symptoms in adolescents [29]. However, research is limited on which processes might promote resilience to the mental health risks of cyberbullying specifically. Such information can inform the development of targeted cyberbullying resilience interventions that can complement efforts to reduce the behaviours entirely.

We focus on modifiable factors that are plausible intervention targets, including any factor influencing resilience from the individual, family, school, community, or environment level. Resilience in mental health is likely the process of many dynamic and interacting biological, psychological, social, and environmental factors that vary across individuals, cultures, and contexts [28]. Despite this complexity and heterogeneity, resilience research has traditionally focussed on a narrow set of variables in cross-sectional studies, such as social support and self-efficacy [30]. Previous studies typically develop theories around a single exposure and examine their associations with mental health outcomes or resilience in isolation despite their likely co-occurrence and collinearity with other exposures [31]. Focussing on individual exposures in isolation simplifies theories of resilience and enables studies to conduct detailed analyses of their association with an outcome to interrogate potential causality. However, mental health research has almost exclusively relied upon these approaches, which risks overlooking potentially important resilience factors and selective reporting. Our study is intended to take a step back from the mental health resilience literature and highlight other potentially overlooked factors that future research may test in a more granular, hypothesis -driven way. The exposure-wide method can essentially act as a systematic hypotheses generation mechanism to inform more traditional studies on narrower or focussed sets of variables.

Population-based cohort studies are rich data resources, and detailed analyses of a few factors exploit the depth but not the breadth of information available. An underutilised complementary approach involves taking a shallow dive into many exposures to systematically identify novel resilience factors and potential intervention targets to inform deeper dives in future studies. Exposure-wide methods examine many exposure-outcome associations within a single study to highlight potential associations of interest [31]. For example, a 2020 study performed an exposure-wide scan of data in the UK Biobank to identify daytime sleep, confiding in others, and television watching time as potentially modifiable mental health risk and resilience factors in adults from a pool of 106 possible exposure variables [32]. However, exposure-wide approaches in adolescents are rare [33, 34], and other approaches considering multiple exposures are informed by systematic reviews, which are subject to publication biases and selective reporting [30]. Many studies also examine resilience as an outcome, despite it being a process of adapting to a specific risk factor, making it more appropriately operationalised as a moderator of a risk factor and outcome association [28].

We aim to address these limitations by conducting an exposure-wide analysis to identify possible resilience factors that might moderate the association between cyberbullying victimisation and psychological distress during adolescence (see Figure 1). We conduct our main analysis in the Millennium Cohort Study (MCS). We perform a replication analysis in the Longitudinal Study of Australian Children (LSAC) to test the robustness of our findings. We first examine longitudinal associations between any potentially modifiable factor measured at baseline and psychological distress at follow-up in MCS to identify possible resilience factors (aim one). We then examine whether any of these possible resilience factors moderate the association between cyberbullying victimisation and psychological distress in the MCS dataset and then examine how many of these findings replicate in a second longitudinal cohort (LSAC) (aim two). Our findings could highlight new intervention targets for future preventative research, which provides added impetus for us to follow co-production principles in this study. Co-production involves conducting research alongside young people with lived experience of mental health conditions, whose perspective can provide uniquely valuable insights and improve the quality of outputs [35]. We incorporate the perspectives of young people with lived experience in this study’s conceptualisation, interpretation, and communication to improve its relevance to the issues we address in youth mental health.

**Figure 1.**
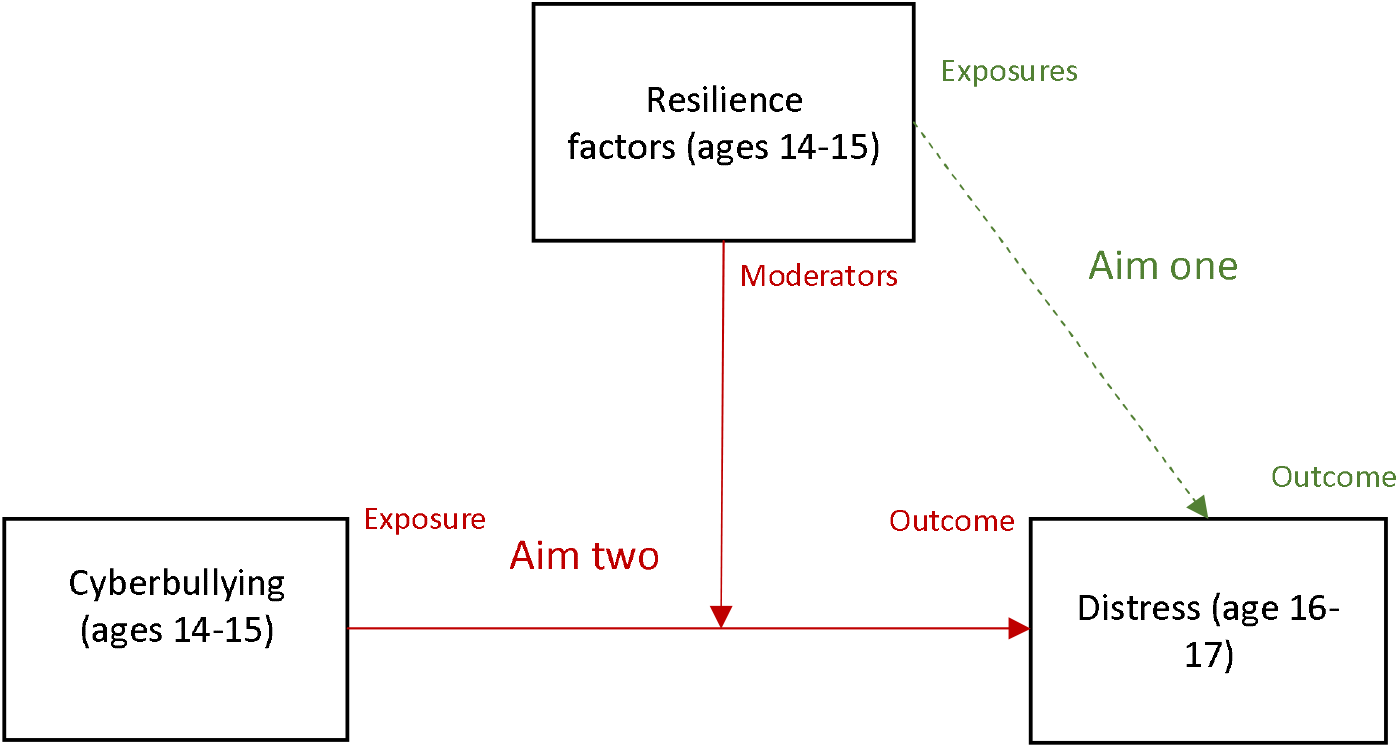
Outline of aims and assumptions.

## 2. Results

### 2.1. Population

There were 9,969 participants (84% of the wave six total) in our main MCS sample who completed baseline measures at ages 14 to 15 and a psychological distress measure at age 17. Mean K-6 scores were 7.30 (standard deviation (SD) = 4.91), and 1,186 (11.91%) had scores ≥14, indicating elevated distress [38].

There were 4,240 participants (50.32% of the total at baseline) in the LSAC dataset, with baseline measures at ages 14 to 15 and a complete outcome score at ages 16 to 17. The mean K-10 scores were 22.05 (SD = 9.05), and sMFQ scores were 7.45 (SD = 7.60). Using the ≥22 and ≥12 for the K-10 and sMFQ, respectively, there were 1,459 (34.41%) people with elevated distress scores. Baseline characteristics for MCS and LSAC are available in Table 1 and 2 of the Supplementary materials.

### 2.2. Aim one: identifying potential resilience factors

We identified 1,466 variables measured at ages 14 to 15 in the MCS dataset that met our inclusion criteria of being plausibly modifiable and unlikely to be a close psychiatric comorbidity of psychological distress. Most of these factors were in the structural domain (43.11%, n = 632), followed by the individual (26.67%, n = 391), family and friends (16.98%, n = 249), learning environment (10.85%, n = 159), and community (2.39%, n = 35) domain. For the first phase of analysis addressing aim one, there was evidence of an exposure-outcome association in 478 of the 1,466 regression models after adjusting for confounders, including sex, birth month, ethnicity, baseline mental health, and parental mental health (see Figure 2). Most of these exposure variables were in the individual domain (38.08%, n = 182), followed by family and friends (24.48%, n = 117), structural (23.85%, n = 114), learning environment (9.62%, n = 46), and community (3.97%, n = 19). These 478 exposure variables form the list of variables we used in the next analysis phase as possible moderators.

**Figure 2.**
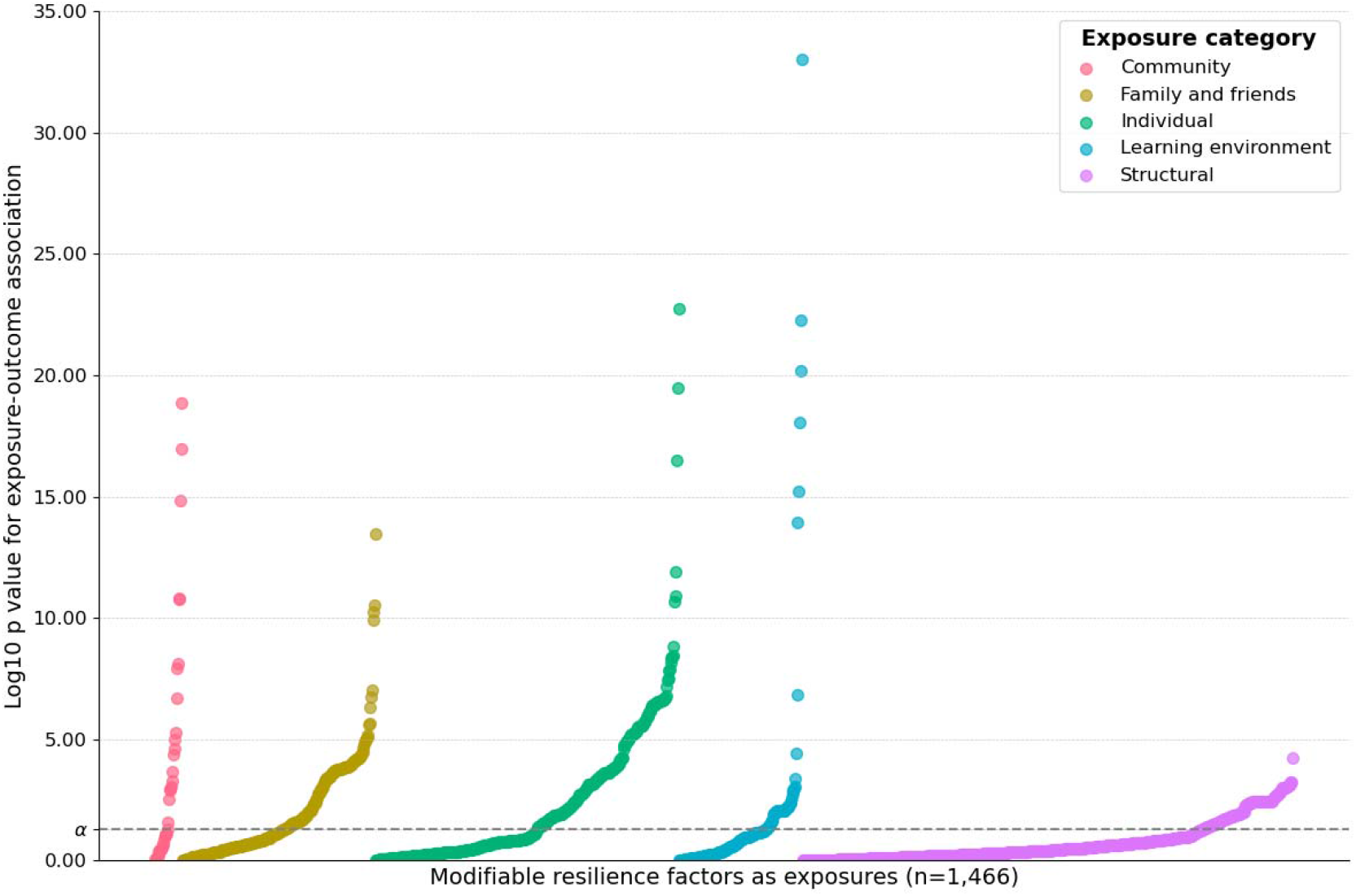
The association of modifiable resilience factors as exposures with psychological distress.

### 2.3. Aim two: investigating resilience factors as moderators

For the second phase of analysis addressing aim two, the exposure was cyberbullying, the outcome was psychological distress, and there were 478 variables from the first analysis phase that we used as moderators. We first confirmed that cyberbullying victimisation was associated with higher psychological distress scores using linear regression (β = 1.98, 95% confidence intervals = 1.76, 2.20, p-value <0.05). We then entered each of the 478 variables from the previous phase of analysis as multiplicative moderators into separate models and adjusted each model for confounders that depended on the domain of the moderator (see Supplementary materials Table 3). There were 31 of 478 variables with evidence of moderation (see Figure 3). Most of these moderators were in the individual domain (54.84%, n = 17, e.g., happiness with friends), then family and friends (19.35%, n = 6, e.g., parental supervision), structural (12.9%, n = 4, e.g., receipt of state benefits), learning environment (9.68%, n = 3, e.g., tiredness at school), and community (3.23%, n = 1, i.e., trust in others). We present the results of the 31 models in Supplementary materials Table 4.

**Figure 3.**
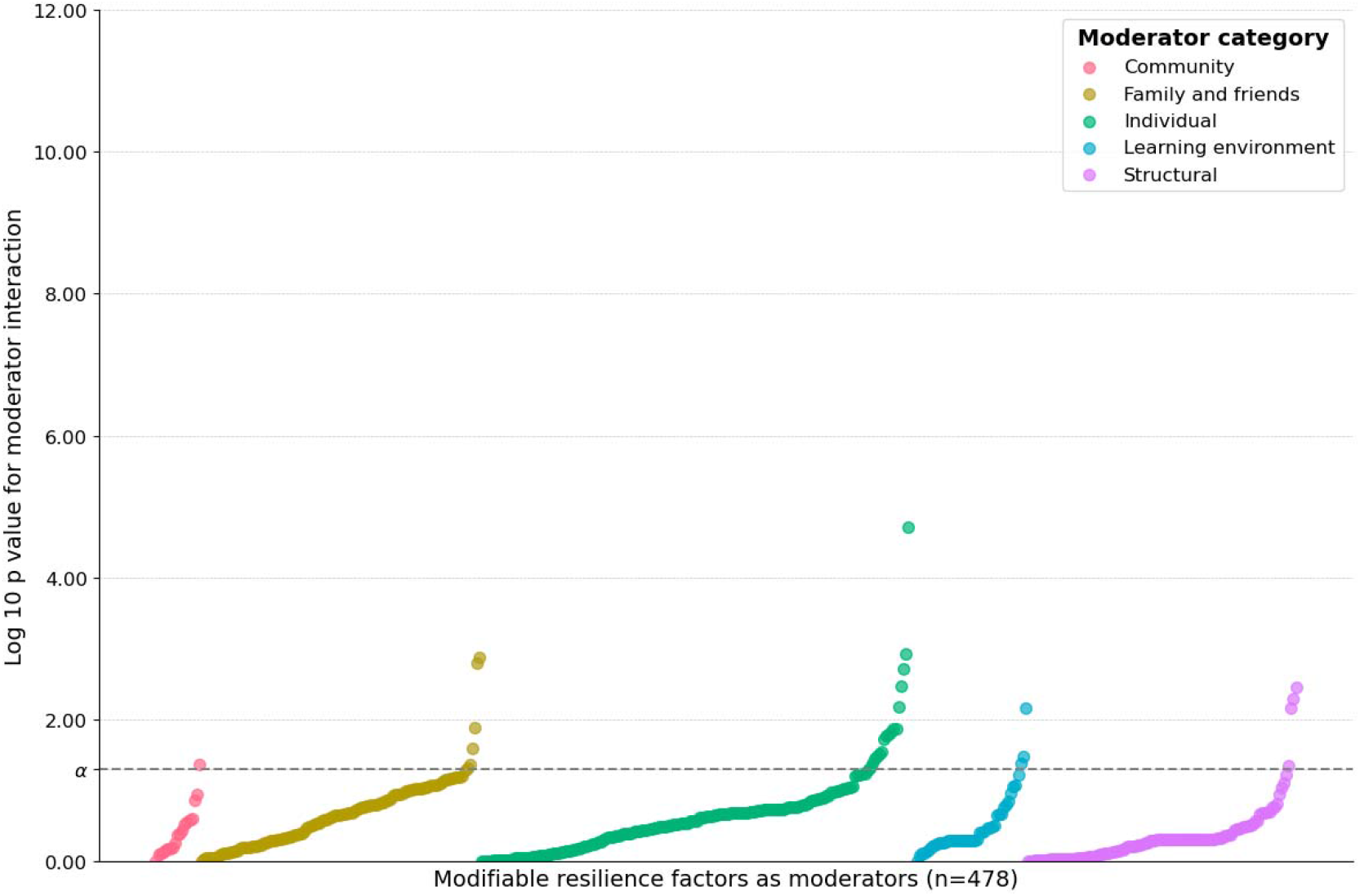
The association of moderators of cyberbullying as the exposure with psychological distress.

### 2.4. Replication analysis

We replicated the second phase of analysis addressing aim two in the LSAC dataset. We found sufficiently comparable moderator variables for 25 of the 31 MCS variables where we found evidence of moderation in the main analysis (see Supplementary materials Table 5). We ran 25 regression models using LSAC variables that match the MCS models in the main analysis and extracted the confidence intervals for the moderator from each model (see Supplementary materials Table 5). We also extracted the moderator confidence intervals for the equivalent MCS models from the main analysis and performed 10,000 simulations per comparison. We calculated the mean estimate and confidence intervals from the 10,000 simulations for each comparison. Supplementary materials Table 6 displays the results of the simulated replication analysis where 15 of the 25 models produced confidence intervals without including zero after the simulations, meeting our criteria for replication. Most of the models that replicated were in the individual (53.34%, n=8, e.g., happiness with life in general), then the family and friends (20%, n = 3, e.g., sexual activities), structural (13.34%, n = 2, e.g., sources of income), and learning environment and (13.34%, n = 2, e.g., misbehaviour in class) domains. Figure 4 includes 15 margin plots showing the association between cyberbullying and psychological distress by each moderator that replicated.

**Figure 4.**
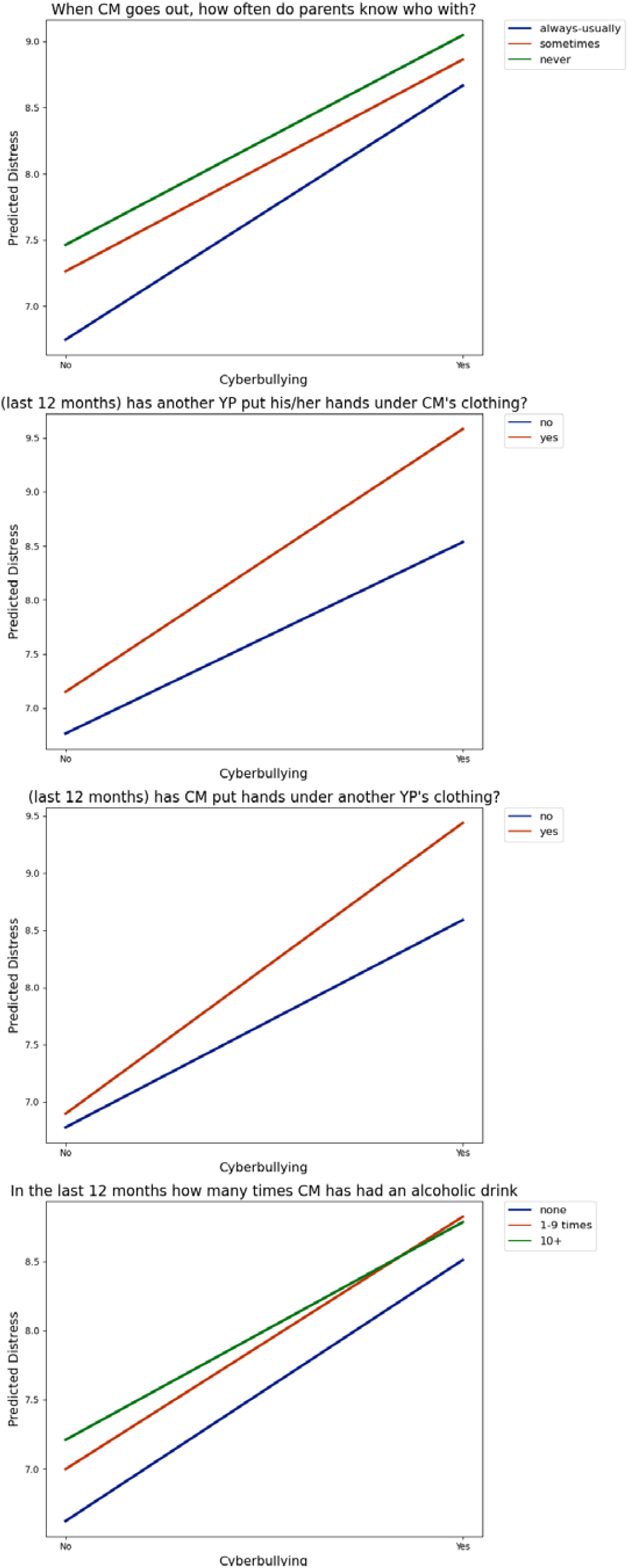
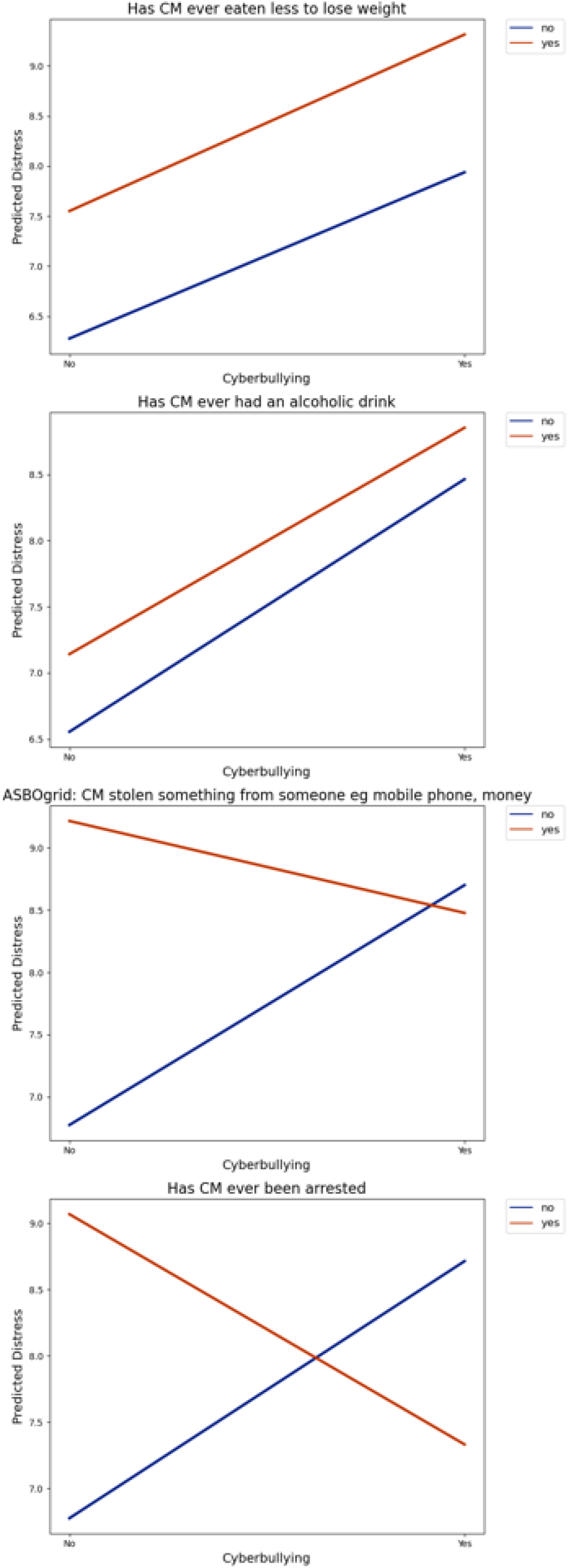
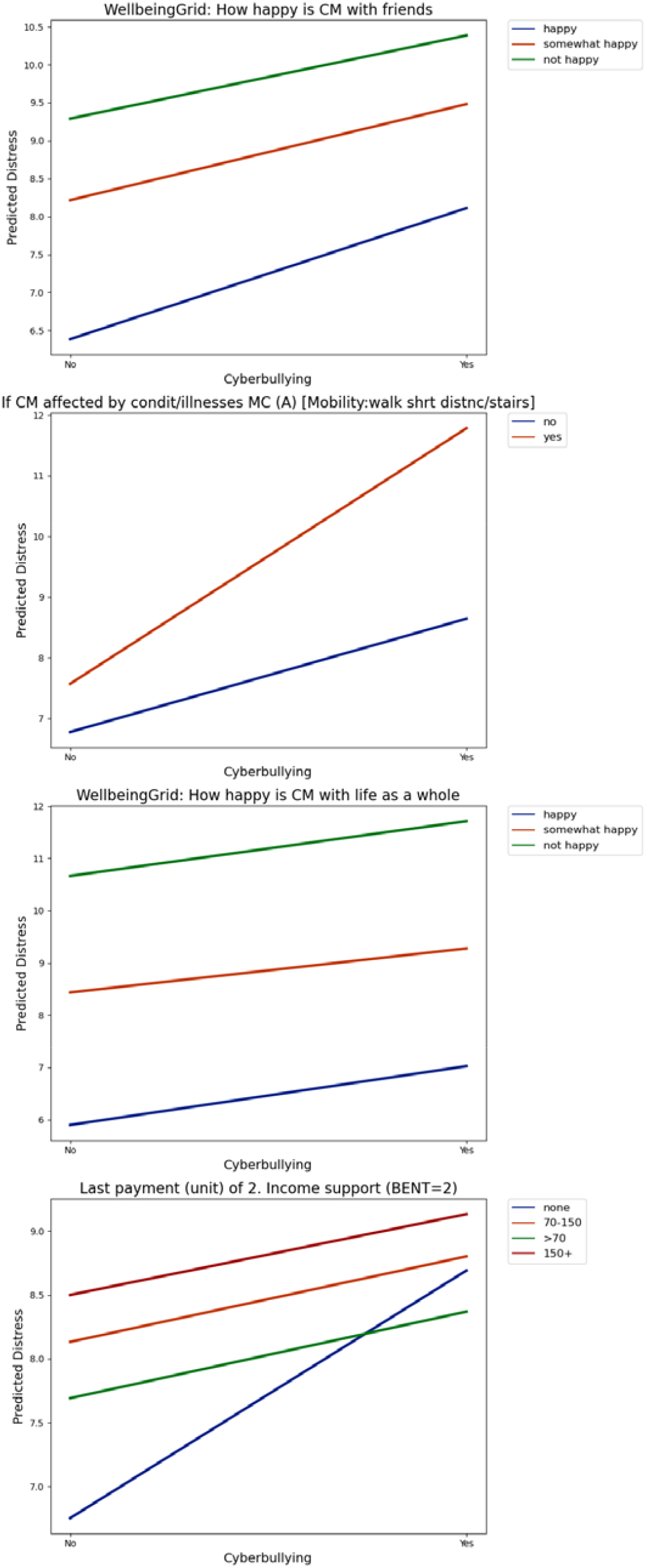
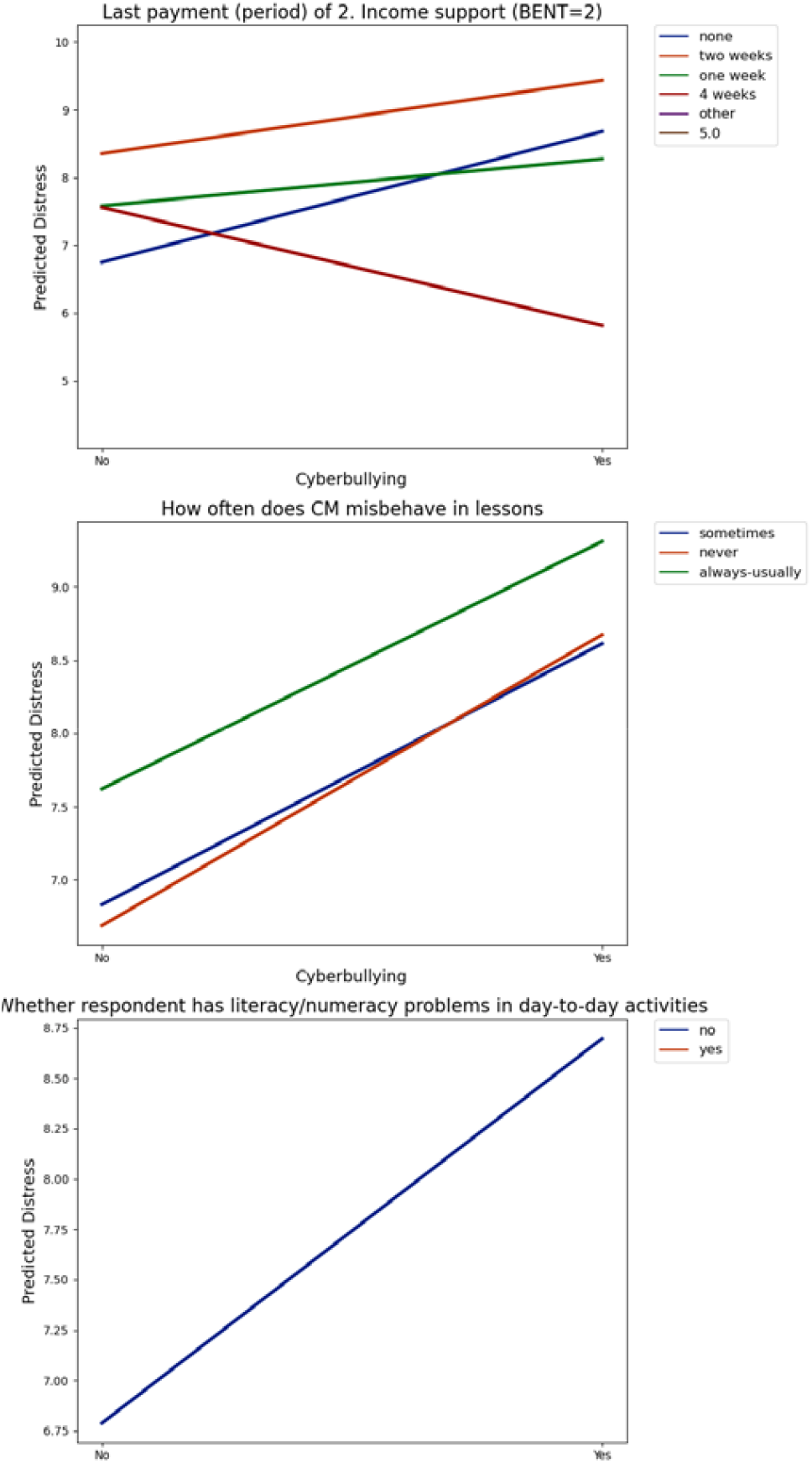
The association of cyberbullying and psychological distress by each moderator that replicated. *Note: these plots only include models that replicated in LSAC and include some additional data cleaning steps to allow for improved visualisation, such as grouping continuous variables into categories.*

### 2.5. Secondary and sensitivity analyses

We repeated the 31 models from the main analyses in the MCS dataset using logistic regression with self-harm as a binary outcome variable. There were 2 of the 31 models with evidence of moderation, whereby the moderator influenced the association between cyberbullying and self-harm (see Supplementary materials Table 7). We also repeated the 31 models from the main analysis in MCS where we used multiple imputations with chained equations to create a complete exposure and confounder variables for each model. The results of these models were comparable with our main analysis (see Supplementary materials Table 8), indicating that sampling bias due to missing data was unlikely to substantially affect our findings.

## 3. Discussion

We leveraged the breadth of data available in two longitudinal national cohorts to identify and examine potentially modifiable resilience factors that could moderate the mental health risks of cyberbullying victimisation in adolescents. Our analysis showed that just under one-third of the modifiable variables measured at age 14 in MCS were associated with psychological distress at age 17. We found that around 6% of these variables moderated the cyberbullying-distress association in the main MCS and LSAC replication analyses, with just over half, reducing the strength of the cyberbullying-distress association. Most of these variables were in the individual domain and were potentially modifiable resilience factors such that they were associated with lower distress scores in people who experienced cyberbullying, including greater happiness with friends or life in general, rarely or never drinking alcohol, and avoiding dieting to lose weight. Family and friends (e.g., sexual activities) was the second largest domain, followed by the learning environment (e.g., literacy or numeracy problems) and structural domains (e.g., receipt of state benefits). Across these domains, avoiding sexual activities, good behaviour during lessons, and parental supervision (e.g. knowing who the child goes out with) were potential resilience factors. Resilience factors related to wellbeing had some of the strongest associations with the lowest error, including happiness with friends and life as a whole. However, several replicated variables were difficult to interpret, such as the income support period.

We identified potential resilience factors across four domains, aligning with the growing recognition of resilience comprising multiple systems [28]. Systematic review evidence suggests that most modifiable resilience factors for young people’s mental health are in the individual domain [49]. We extend existing findings by showing the importance of individual domain factors when considering all measured variables in our dataset rather than only considering the existing literature, which is subject to publication bias and selective reporting. Individual domain factors may be more amenable to direct interventions (e.g., alcohol use and parental supervision) than some other domains, such as income (structural).

There is some overlap between the resilience factors we identified in this study and those included in two prominent frameworks of multisystem resilience by Masten *et al*. [50] and Ungar *et al*. [51]. For example, alcohol consumption and dieting behaviours relate to Ungar’s inclusion of experiences of control and efficacy, which includes the capacity for self-care. In our results, dieting behaviours had a marginally higher impact as a moderator. Both frameworks include attachment and relationship with family and peers, which relate to our findings that happiness with friends and parental knowledge of who the young person goes out with are possible resilience factors. Our findings suggest that happiness with friends is a stronger moderator than parental supervision. These factors indicate high-quality relationships with friends and family that could create a supportive environment that fosters resilience to cyberbullying. They could also reflect adolescents around this age prioritising forming friendships over parental relations. However, we found no community domain factors replicated in our analysis, which are prominent in both frameworks, such as having spiritual and cultural belief systems. We also found evidence for possible resilience factors outside these frameworks, such as specific sexual activities.

These differences may be due to our examination of resilience as a moderator within the context of cyberbullying and psychological distress rather than as an outcome [28]. For example, several studies highlight social support and positive family climate as resilience factors moderating the association between mental health outcomes and risk factors, including parental psychopathology and childhood adversity [30, 49]. Our findings that happiness with friends and parental knowledge of who the young person goes out with as possible resilience factors suggest that these findings may extend to cyberbullying as a risk factor. Parental monitoring has also been shown elsewhere to be associated with reduced depression, anxiety, and self-harm in adolescents and could be a promising intervention target [52]. However, these studies did not examine health behaviours that we identified as possible risk factors, such as alcohol use and dieting. Identifying such factors of interest that were not included in other studies is an important aspect of implementing these exposure-wide methods in large cohort studies.

This study is a comprehensive examination of 1000s of variables to systematically identify modifiable resilience factors that could potentially reduce the risk of psychological distress after cyberbullying victimisation in adolescents. These exposure-wide approaches are an underutilised method in epidemiology that can inform and complement studies that focus on a narrower set or single variables. Scanning large numbers of variables across multiple domains while minimising the reliance on published literature is particularly suitable for examining the complex and dynamic processes of resilience and reduces the risk of missing important influences. We increased the robustness of our findings by replicating our analysis in a second cohort and followed our pre-registered protocol (https://doi.org/10.17605/OSF.IO/UPZTS) to increase transparency. We also increased the relevance and applicability of our research by incorporating lived experience perspectives, engaging with a youth advisory group and involving a young person with lived experience in our core study team and co-authoring the manuscript.

There are also several limitations to our study, such as the exposure being a single-item, binary response measure that may not capture the full extent of cyberbullying in MCS participants. However, around 26% of young people in our study reported being victims of cyberbullying, which is higher than estimates in other studies using more comprehensive measures in a similar sample [53]. Dichotomising the exposure also meant that we grouped participants who experienced different frequencies of bullying, which could influence what moderators might influence the association with distress. We could also not exclude the possibility of missing a potential resilience factor as we were restricted to variables measured in MCS and LSAC cohorts. The breadth of variables in these cohorts still enabled us to take a comprehensive and systematic approach that included over 1000 potential resilience factors. However, handling such large numbers of variables means we do not perform the detailed individual variable management and coding steps of more focussed epidemiological studies. The large number of models also meant that we relied on p values as a crude but efficient metric of evidence for an association in the first phase of analysis, which may have introduced false positives or negatives. However, we used a more sophisticated bootstrapping method to determine whether our main findings were replicated, which reduced the likelihood of error in our final list. The longitudinal nature of the MCS data means that there is attrition between waves, which can cause sampling biases by disproportionately affecting certain groups, such as those in rented accommodation or from Pakistani or Bangladeshi backgrounds [54].

The exposure-wide approach taken here is a shallow but wide approach that gives less attention to the nuances of each variable and its mechanisms than a deep but narrow approach. A consequence is that some variables in the final results were highly skewed, such as literacy and numeracy problems (yes = 2, no = 9,412) or being arrested (yes = 83, no = 9572). Others were difficult to interpret, such as the period of income support payment. Some of these variables are likely statistical artifacts, but we chose against using corrections for multiple comparisons to avoid suppressing weak but potentially viable findings, as is common in epidemiological research [47].

We tested the robustness of our findings by replicating the MCS findings in the LSAC, a cohort with several similar properties, including age ranges, cohort periods, and measures. However, we could not find suitable comparison variables for six MCS variables in LSAC, and there were several cases where the variables only loosely matched. For example, we used an LSAC variable indicating whether the participant had ever had sex to approximate the MCS variables on whether the participant had touched someone or been touched by someone under their clothing. The exposure variable was also in a different format to MCS as it specified different types and mediums for cyberbullying, which required collating into a binary variable for comparability. The cross-sequential design of the LSAC also meant that a subsection of the cohort had a different measures (SMFQ) as their outcome rather than the K-6 distress scale. The scales aim to capture similar concepts over different timeframes (2 weeks vs 1 month), and we were able to standardise and combine them, but this may have affected the replicability of our findings. Our LSAC sample was also less than half the size of the MCS sample. This could introduce additional sampling biases and mean that a lack of statistical power could explain why some variables failed to replicate. However, our bootstrapping method was less sensitive to smaller sample sizes than other metrics, such as p values. Some of these limitations are difficult to avoid when conducting research across multiple independent cohorts, but we believe the results are ultimately more robust from examining the replication of our main findings in a second cohort.

We used an exposure-wide approach to comprehensively and systematically identify possible resilience factors that moderate the association between cyberbullying victimisation and psychological distress in adolescents. We found 15 modifiable factors that moderated the cyberbullying-distress association in the MCS and LSAC datasets. These resilience factors were mostly in the individual domain (e.g., low alcohol use), and some of the strongest moderators with the lowest error were related to wellbeing, including happiness with friends and life in general. Our exposure-wide approach aimed to conduct a shallow dive into many associations to identify resilience factors that are potential intervention targets to reduce the mental health risks of cyberbullying and require future research to conduct deeper dives to better understand the effects of these factors and their mechanisms. For example, future research could explore reducing the mental health risks of cyberbullying by promoting specific aspects of wellbeing identified in this study, such as relating to friends and life in general. Another approach could be to examine how wellbeing interacts with other possible resilience factors from this study to influence mental health risks, such as parental supervision or dieting. Researchers could explore using exposure-wide designs as a means of systematic hypothesis generation to inform more traditional epidemiological studies that focus more deeply on a narrow set of variables. Future research should also explore exposure-wide methods in other cohorts to examine how resilience may vary according to age, culture, socioeconomic groups, and other relevant influences. The complementary use of these methods can reduce the risk of overlooking potentially important associations and selective reporting while embracing the complexity of dynamic, multisystem concepts like resilience. Our findings will directly inform further research within the wider Digital Youth research programme (https://digitalyouth.ac.uk/) on certain resilience factors and interventions to reduce online harms in young people.

## 4. Methods

### 4.1. Population

We used data from adolescents in two population-based cohort studies, including the United Kingdom-based Millennium Cohort Study (MCS, n = 19,517) and the Australia-based Longitudinal Study of Australian Children (LSAC, n = 10,090). The MCS data formed our primary analytic sample, and the LSAC data was used for the purpose of replicating the main analysis as described below. The MCS consisted of individuals born between September 2000 and January 2002 across the UK using a stratified cluster design and includes seven waves of data collection via surveys, biological samples, and other methods. Full details on the design, methodology, data availability, and other aspects of the cohort are available at https://cls.ucl.ac.uk/cls-studies/millennium-cohort-study/. We used data from MCS participants aged 14 to 15 at wave six (2014 to 2015, n = 11,872, 61.3% of original sample) as our baseline and aged 17 at wave seven (2018 to 2019, n = 10,757, 55.1% of original sample) as our follow-up point. We chose this age range as it represents a critical period for the development of mental health problems [36]. The National Research Ethics Service (NRES) Research Ethics Committee (REC) North East – York (REC ref: 17/NE/0341) and National Research Ethics Service (NRES) Research Ethics Committee (REC) London – Central (REC ref: 13/LO/1786) provided ethical approval for the MCS, and no additional ethics approval is required for this analysis.

The LSAC is a cross-sequential national study that includes a Birth cohort (B cohort) of 5,107 children born between 2003 and 2004 and the Kindergarten cohort (K cohort) of 4,983 children enrolled at ages four to five but born in 1999 to 2000. The study includes nine waves of data collection and used a two-stage, clustered sampling design, which is described in full with other relevant information on the data, design, and sample characteristics on the study website (https://growingupinaustralia.gov.au/). We used data from children aged 14 to 15 at waves eight (B cohort, 2018, n = 4,030) and six (K cohort, 2014, n = 4,395) as our baseline (combined n =8,425). Our follow-up period was around ages 16 to 17 at waves 9C1 (B cohort, 2020, n = 1,595) and seven (K cohort, 2016, n = 3,537). The cross-sequential design and comparable measures of LSAC allow us to combine the B and K cohorts into a single sample. The Australian Institute of Family Studies Ethics Committee provided ethical approval for the LSAC and no additional ethics approval is required for this analysis.

Our analytical sample for this study included children who participated in the baseline assessments at ages 14 to 15 and had a completed outcome measure at follow-up ages 16 to 17 in MCS (primary cohort, n = 9,969) and LSAC (replication cohort, 4,240). The participant ages and data collection dates are comparable across MCS and LSAC at our baseline and follow-up points, and we present other sociodemographic characteristics in the results section. Before conducting this study, we pre-registered our protocol on the Open Science Framework (https://doi.org/10.17605/OSF.IO/UPZTS).

### 4.2. Outcome(s)

Our primary outcome was psychological distress at age 17 (wave seven) in the MCS. The cohort used a six-item Kessler Psychological Distress Scale (K-6), which measures distress during the past month on a five-point Likert scale, with total scores ranging from 0 (low) to 24 (high) [37]. We selected the K-6 scale, a brief and well-validated measure of common mental health symptoms that performs similarly to other condition-specific scales for depression and anxiety screening, such as the Patient Health Questionnaire and Generalised Anxiety Disorder scales [38].

We used the ten-item Kessler Psychological Distress Scale (K-10) from the B cohort and short Moods and Feelings Questionnaire (sMFQ) from the K cohort at ages 16 to 17 in LSAC for the replication outcome [37]. The K-10 measures psychological distress with properties similar to the K-6 version, with additional items for depressed mood, agitation, fatigue, and nervousness. It uses a five-point Likert scale with scores ranging from 10 (low) to 50 (high). The sMFQ measures depressive symptoms using 13 items that cover the past two weeks on a three-point Likert scale, with total scores ranging from 0 to 26 [39]. We used the sMFQ because the K-10 measure was unavailable at the comparable follow-up time point in the K cohort. The K-10 and sMFQ are comparable as they target similar aspects of distress. We use z scores to standardise the K-10 and sMFQ in LSAC to combine them into a single outcome score in the replication models.

We entered the scales as continuous outcomes to more closely represent the reality of distress occurring on a continuum rather than a discrete binary outcome, such as a clinical diagnosis. However, we provide data on binary outcomes for descriptive purposes using the established thresholds for clinical relevance of ≥14 for K-6 [38], ⍰0≥22 for K-10, and ≥12 for sMFQ [40].

Our secondary analyses also included self-reported self-harm behaviour as a secondary outcome at ages 16 to 17. The MCS and LSAC measure self-harm with binary ‘yes’ or ‘no’ responses to a question about intentional self-harm. The item in MCS is: *During the last year, have you hurt yourself on purpose in any of the following ways? Cut or stabbed, burned, bruised or pinched, overdose, pulled out hair, other. The item in LSAC is: During the past 12 months have you hurt yourself on purpose in any way (i.e., by taking an overdose of pills, or by cutting or burning yourself)?*

### 4.3. Exposures and moderators

We initially considered modifiable resilience factors as exposure variables during the first phase of analysis addressing aim one (see Figure 1). We identify these resilience factors by manually curating a list from all measured variables at age 14 in MCS that are plausibly modifiable at an individual, family, school, community, or structural level and are unlikely to be a close proxy for psychiatric comorbidity of psychological distress, such as Strengths and Difficulties Questionnaire scores. All variables meeting these two criteria are included as exposure variables in the aim one analysis.

Given that this is an exploratory phase of analysis that includes many variables of different types, we conducted minimal data cleaning or reformatting before analysis. One author (AK) created the list from the data dictionary, a second author (RM) independently verified the list, and a third author (PP) mediated any discrepancies between the lists of each author. We organised each factor in the list into domains for descriptive purposes using Public Health Scotland’s Children and Young People’s Mental Health Indicators framework, which includes individual, family and friends, learning environment, community, and structural domains [41]. Public Health Scotland developed this framework to describe population mental health outcomes and determinants through consultation with domain experts and community adults, children, and young people. Given the high quantity of exposure variables we will include in the analysis, these domains aid in interpreting the results. We retain all potentially modifiable resilience factors longitudinally associated with distress scores following the analytical plan outlined below to derive a final list.

Cyberbullying victimisation is the exposure variable in the second phase of analysis addressing aim two (see Figure 1). The MCS measures cyberbullying as a six-point self-reported response scale (Never to Most days) to the question: *How often have other children sent you unwanted or nasty emails, texts or messages or posted something nasty about you on a website?* We created a binary exposure whereby we considered the responses ‘Most days’, ‘About once a week’, ‘About once a month’, and ‘Every few months’ as experiencing cyberbullying (1) and ‘Less often’ and ‘Never’ as not experiencing cyberbullying (0). We use this single-item response as an indicator of cyberbullying victimisation as in other studies [42]. The LSAC measured cyberbullying using items from the School Climate Bullying Scale and Edinburgh Study of Youth Transitions and Crime modified for an Australian context, which included a list of 14 potential acts of online bullying in the past 30 days [43, 44]. We created a binary exposure variable whereby participants had experienced any of the 14 forms of online bullying as a victim (1) or none (0), as in previous studies [45].

The second phase of analysis also included moderators that potentially influence the association between cyberbullying (exposure) and psychological distress (outcome) in the MCS (see Figure 1). We identified these possible moderators from the final list in aim one and made no adjustments to their values for the main analysis, i.e., no further cleaning or reformatting. However, the replication analysis requires finding comparable variables in the LSAC dataset for each MCS moderator in the final list, where possible. See Supplementary materials Table 3 for details on the MCS variables matched with LSAC variables and grading of their comparability. It was also necessary to harmonise the variables across the MCS and LSAC datasets to ensure meaningful comparisons during replication. This harmonisation includes reformatting or cleaning certain variables to make them comparable, such as recoding a variable from having five to three categorical responses. We made such adjustments only where necessary and only to the LSAC dataset to preserve the main MCS dataset findings where possible. All adjustments are visible in the code that we provide with this manuscript.

### 4.4. Covariates

All models aim one are minimally adjusted for confounding variables unlikely to be on the exposure-outcome pathway (see Figure 1), including sex (as indicated at baseline), ethnicity (as indicated at baseline), birth month, and baseline parental mental health (to approximate genetic mental health risk, K-6 scale). We also adjust for baseline mental health symptoms (sMFQ) to reduce the risk of reverse causation.

We adjusted aim two models for the same variables as in aim one (confounders of the aim two moderator-outcome pathway in Figure 1) and baseline covariates that may confound the association between cyberbullying and psychological distress at follow-up (aim one path of Figure 1). These confounders include sex, ethnicity, parental mental health, birth month, household income (quintiles), parental education (highest parental National Vocational Qualification), and household family composition (one or two parents at home). We also adjusted for additional variables that may confound the association between each resilience factor (moderator) and psychological distress (outcome). The exact composition of these additional confounders varies in each model depending on the domain of the resilience factor (see Supplementary materials Table 3 for specific confounders in each domain) but include smoking frequency (including e-cigarettes), alcohol use in the last four weeks, disability (in MCS, a condition that limits daily activity), social support (Short Social Provisions Scale), self-rated general health level, and area-level deprivation (Index of Multiple Deprivation).

### 4.5. Statistical analysis

For aim one, we used an exposure-wide approach [46] by entering each baseline resilience factor as an exposure into a separate univariate linear regression with psychological distress at follow-up. The initial models only include complete cases per exposure. We adjusted each model for potential confounders that are unlikely to be on the exposure-outcome causal pathway as described above. We used the statistical threshold of *p* < 0.05 as an indicator of evidence for an exposure-outcome association after confounder adjustments and include any exposure within this threshold in our list of modifiable resilience factors for the next phase of analysis addressing aim two. P values were an efficient way to scan many exposure-outcome associations for signal amongst the noise, and we did not adjust for multiple comparisons to avoid suppressing that signal.

In separate linear regression models, we examined each modifiable resilience factor from our aim one list as a moderator of the association between cyberbullying (exposure) and psychological distress (outcome) for aim two. We adjusted these models for the confounders from the aim one model plus additional variables that may confound the exposure-outcome and domain-specific moderator-outcome pathway following the strategy we outlined in the previous section. A list of confounding variables for each model according to the moderator domain is available in Supplementary materials Table 3. We entered the moderator into each model as a multiplicative interaction term with the exposure and used a statistical threshold of p < 0.05 as an indication of evidence for moderation. We chose against correcting for multiple comparisons here as such procedures reduce the risk of false positives (type 1 error) at the expense of increasing the risk of false negatives (type 2 error), which is a particular problem in epidemiological research where datasets typically contain small effects with substantial noise [47]. Therefore, multiple corrections would suppress potentially real effects and contrast with our aims of highlighting associations of interest that the current literature may have overlooked for future studies to examine in detail [48]. We are mitigating the increased risk of type 1 error by replicating our findings in a second cohort (described below) and highlighting only replicated findings as potential candidates for future studies to investigate their association with psychological distress in more detail.

### 4.6. Replication

The next step was the replication phase of analysis, where we aimed to replicate any MCS model that shows evidence of moderation in the LSAC dataset as closely as possible. Supplementary materials Table 3 shows the original MCS confounder and moderator variables alongside the LSAC equivalent we used in the replication analysis. Using their LSAC equivalents, we repeated the models from the previous step of analysis, where there was evidence of moderation in MCS.

We used a bootstrapping method to determine whether a model in the MCS dataset replicates in the LSAC dataset. We chose this as a stable and robust method that avoids making assumptions about the data distribution, is less reliant on sample size, and better accounts for effect size and direction than using p values alone to determine replication. The method involves extracting the 95% confidence intervals for the exposure-moderator interaction term from each model in the MCS and LSAC datasets. We randomly draw a number between the intervals for MCS and LSAC to calculate the difference between them and repeat this process 10,000 times for each set of models. We calculated the mean difference with 95% confidence intervals across the 10,000 simulations for each set of models. We then checked whether the confidence intervals for each model included 0, which we considered as an indicator of non-significance. We classified the finding in the MCS dataset to have replicated in the LSAC dataset for any set of models where the confidence intervals for the mean difference following the 10,000 simulations do not cross 0. Our final list of resilience factors included only those showing evidence of moderation in MCS and LSAC, i.e., the replicated variables.

### 4.7. Secondary and sensitivity analyses

In a secondary analysis, we repeated aim two models (investigating resilience factors as moderators) with a dichotomised self-harm outcome to examine the applicability of these assumptions to another mental health outcome. In a sensitivity analysis, we repeated aim two models using a dataset with multiple imputations via chained equations and all other variables of interest to impute missing data (iterations = 30) to assess the possible impact of bias in our results due to missing data.

### 4.8. Co-production

This study was part of the UKRI-funded Digital Youth (https://digitalyouth.ac.uk/) programme, which aims to bring together leading researchers and young people with experience of mental health problems (Sprouting Minds) to understand youth mental health and wellbeing within the context of digital technologies and co-produce and co-design practical preventive and therapeutic interventions. The programme holds regular workshops, talks, research days, and events to share learning and incorporate young people’s perspectives into the research projects at different stages. For example, our study team had a workshop with members of the Digital Youth Young Person’s Advisory Group ‘Sprouting Minds’ to discuss mental health resilience factors during the early phases of this study, focusing on identifying factors potentially missing from our datasets. We also invited a young person from Sprouting Minds to join our core research team to be more directly involved in the study and co-author this paper (YR).

## Data Availability

All data in the manuscript available at: https://cls.ucl.ac.uk/cls-studies/millennium-cohort-study/ and https://growingupinaustralia.gov.au/data-and-documentation

## Data and code availability

The MCS dataset is available through the UK Data Service (https://ukdataservice.ac.uk/) and LSAC through the Australian Data Archive DSS Longitudinal Studies (https://dataverse.ada.edu.au/dataverse/DSSLongitudinalStudies). The code for this analysis will be available through GitHub.

## Acknowledgments

The authors acknowledge the support of the UK Research and Innovation (UKRI) Digital Youth Programme award [MRC project reference MR/W002450/1], which is part of the AHRC/ESRC/MRC Adolescence, Mental Health and the Developing Mind programme. We are grateful to Sprouting Minds and the wider Digital Youth Programme for their involvement and support for the study. We are also grateful to all participants and staff involved in MCS and LSAC.

## Author contributions

AK, YK, PP conceptualised the study and all authors contributed to the design. AK analysed the data. AK, YK, PP, RM, YR, KC, CH, and ET contributed to the interpretation and write-up of the study and approve it for submission.

